# Emerging pathogens associated with acute respiratory infections in children before, during and after the COVID-19 pandemic in Hanoi, Vietnam

**DOI:** 10.1101/2024.11.02.24316627

**Authors:** Nhan Thi Ho, Hang Thi Thuy Nguyen, Ha Thi Hoang, Dung Van Pham, Quang Ngoc Nguyen, Huong Thi Minh Le, An Nhat Pham, Phuong Mai Doan

## Abstract

The COVID-19 pandemic has caused changes in respiratory infectious disease patterns. We analyzed de-identified data of microbiology assays of nasopharyngeal samples and blood samples of children with acute respiratory infection (ARI) visited Vinmec Times City International hospital in Hanoi, Vietnam from January 2019 to December 2023 to examine the overall pattern of viral and bacterial emerging pathogens associated with ARI in children before, during and after the COVID-19 pandemic. The data was aggregated by month and time series analysis and visualization was done. Bacterial Polymerase Chain Reaction (PCR) panel was done for 4125 samples, *Mycoplasma pneumonia* (MP) IgM was done for 5049 samples, bacterial culture was done for 10280 samples and viral PCR or rapid test was done for 42041 samples. After the COVID-19 pandemic, *Haemophilus influenzae* (HI) and *Streptococcus pneumoniae* (SP) have re-emerged as epidemic pathogens associated with lower respiratory tract infection (LRI). *Influenza type A* and *type B* have reestablished regular cycles of peaks in winter-spring months after an early rebound together with an unprecedented newly emergence of *Human Adenovirus* (HAdV) soon after the relief of COVID-19 restriction. Late after the COVID-19 pandemic, from middle of 2023, atypical pneumonia pathogen *Mycoplasma pneumonia* (MP) has emerged remarkably and becoming epidemic and there was also a small, brief emergence of *Chlamydophila pneumoniae* (CP) infection. Our data is useful to understand the influence of COVID-19 pandemic on the emergence or re-emergence of viral and bacterial respiratory infection pathogens in children and is useful for disease surveillance and public health intervention.

## INTRODUCTION

The COVID-19 pandemic and related restricted policies have caused changes in respiratory infectious disease patterns (1–3). The incidence of most of respiratory infections apart from COVID-19 decreased during the COVID-19 pandemic due to restricted measures (2). After easing COVID-19 related restricted measures, the sudden increase of exposure in the community whose immunity might be waned during the prolonged no-contact quarantine period might lead to irregular respiratory infection pattern (1). There have been some reports regarding the rebound of viral pathogens such as Influenza (4,5) and bacterial pathogens such as Mycoplasma (6) after the COVID-19 pandemic. We have previously reported molecular data of case series of *Human Adenovirus* (HAdV) outbreak in 2022 (7) and *Mycoplasma pneumonia* (MP) outbreak in 2023 (8) in Hanoi, Vietnam. In this report, we describe the overall pattern of viral and bacterial emerging pathogens associated with acute respiratory infections (ARI) in children before, during and after the COVID-19 pandemic in Hanoi, Vietnam to understand the influence of COVID-19 pandemic on the emergence or re-emergence of other respiratory infection pathogens in children.

## METHODS

Between August 27, 2024 and September 30, 2024, we accessed and analyzed deidentified data of microbiology assays of nasopharyngeal samples and blood samples of children with ARI visited Vinmec Times City International hospital in Hanoi, Vietnam from January 1, 2019 to December 31, 2023. (We had no access to information that could identify individual participants during or after data collection). The microbiology assays included: **(A)** Allplex Respiratory Panel 4 using real-time Polymerase Chain Reaction (PCR) of nasopharyngeal samples for the detection of 7 bacteria causing respiratory tract infections including *Bordetella parapertussis* (BPP), *Bordetella pertussis* (BP), *Chlamydophila pneumoniae* (CP), *Haemophilus influenzae* (HI), *Legionella pneumophila* (LP), *Mycoplasma pneumoniae* (MP), *Streptococcus pneumoniae* (SP); **(B)** Blood IgM serology test for *Mycoplasma pneumonia*; **(C)** Bacterial culture of nasopharyngeal samples; **(D)** Polymerase Chain Reaction (PCR) or rapid test for *Influenza type A* (Flu A), *Influenza type B* (Flu B), *Respiratory Syncytial Virus* (RSV) and Allplex Respiratory Panel 2 using one-step real-time RT-PCR of nasopharyngeal samples for the detection of 7 viruses causing respiratory tract infections including *Human Adenovirus* (HAdV), *Enterovirus* (HEV), *Metapneumovirus* (MPV), *Parainfluenza virus* 1,2,3 4 (PIV1,2,3,4).

Available clinical data accompanied these microbiology assay data included diagnosis, age, and gender. Patient diagnoses were classified into two groups: upper respiratory infections (URI) and lower respiratory infections (LRI). All patients included in the analysis were ≤15 years old and were from Hanoi or surrounding provinces.

The data were aggregated by months and time series plots were used to visualize the data by month from January 2019 to December 2023. The number of patients tested and the number of patients positive for each pathogen were displayed. For patient characteristics, mean age with standard error, percentage of male, and percentage of diagnosis category LRI were plotted. Co-detection (≥ 2 pathogens detected in a patient) was also described.

Patient characteristics were compared between groups using Kruskal Wallis’s test for continuous variables and Chi-square test for categorical variables.

## RESULTS

Patient characteristics were summarized in **Table 1** and plotted as time series in **Figure 1**. Males accounted for ∼60% of ARI patients for all pathogens. Most bacteria associated ARI patients had lower respiratory infections (LRI) whereas most virus associated ARI patients (except RSV) had upper respiratory infections (URI). Most ARI patients were young (average age ∼3 years old or younger) except those infected with MP, Flu A, and Flu B were a bit older (average age ∼ 5 years old).

**Table 1.**
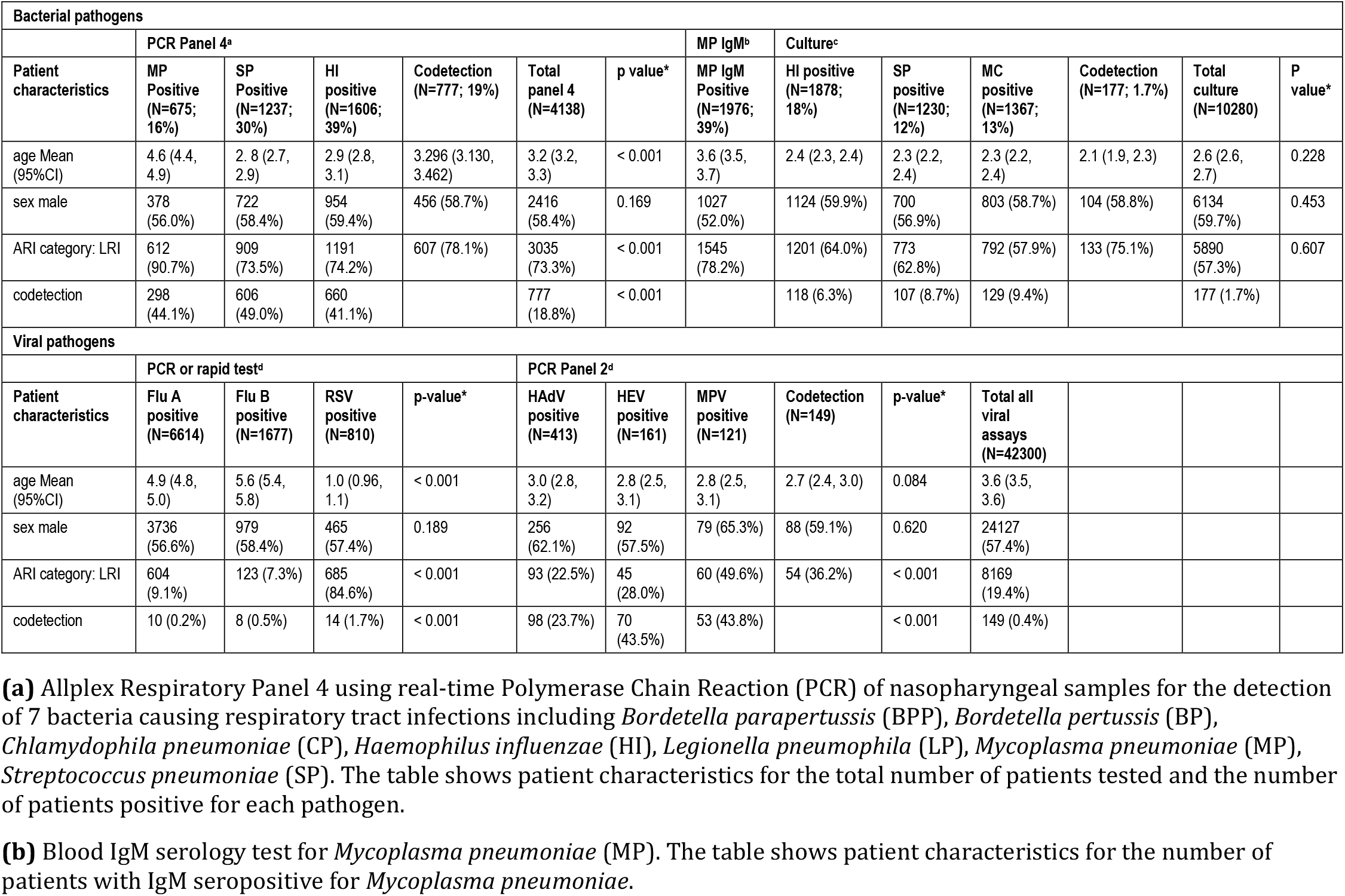
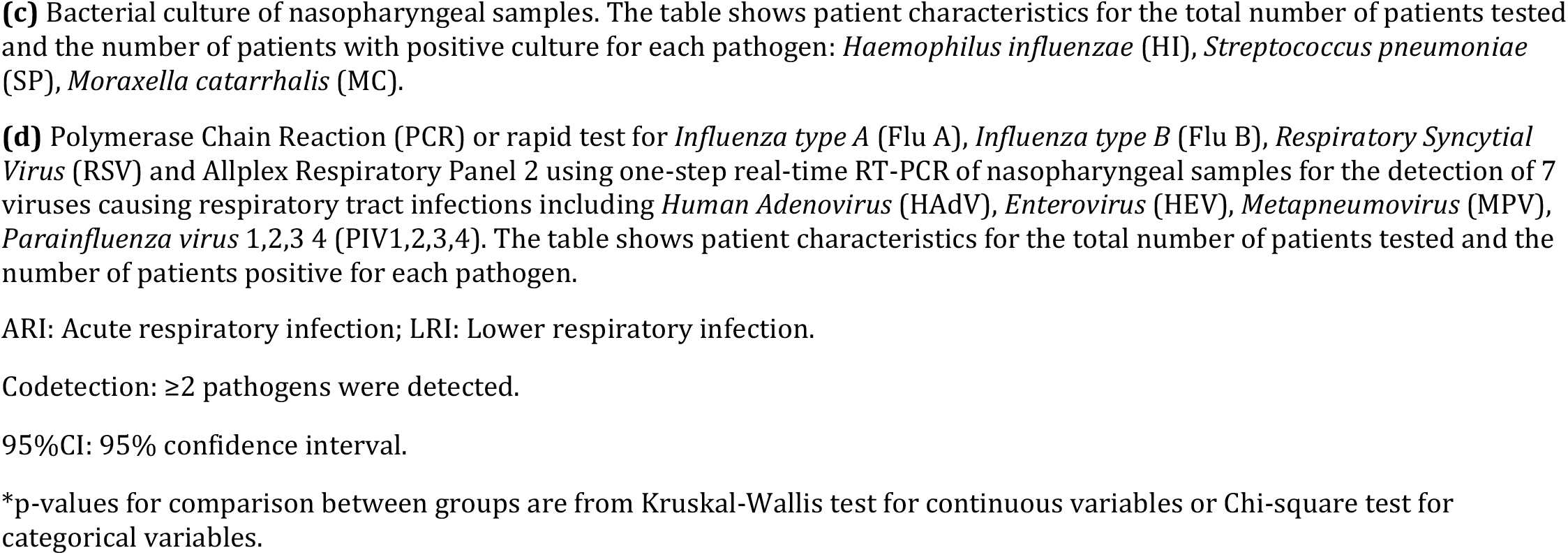
Summary of patient characteristics.

| Bacterial pathogens |  |  |  |  |  |  |  |  |  |  |  |  |  |
| --- | --- | --- | --- | --- | --- | --- | --- | --- | --- | --- | --- | --- | --- |
|  | PCR Panel 4<sup>a</sup> |  |  |  |  |  | MP IgM<sup>b</sup> | Culture<sup>c</sup> |  |  |  |  |  |
| Patient characteristics | MP Positive (N=675; 16%) | SP Positive (N=1237; 30%) | HI positive (N=1606; 39%) | Codetection (N=777; 19%) | Total panel 4 (N=4138) | p value* | MP IgM Positive (N=1976; 39%) | HI positive (N=1878; 18%) | SP positive (N=1230; 12%) | MC positive (N=1367; 13%) | Codetection (N=177; 1.7%) | Total culture (N=10280) | P value* |
| age Mean (95%CI) | 4.6 (4.4, 4.9) | 2.8 (2.7, 2.9) | 2.9 (2.8, 3.1) | 3.296 (3.130, 3.462) | 3.2 (3.2, 3.3) | < 0.001 | 3.6 (3.5, 3.7) | 2.4 (2.3, 2.4) | 2.3 (2.2, 2.4) | 2.3 (2.2, 2.4) | 2.1 (1.9, 2.3) | 2.6 (2.6, 2.7) | 0.228 |
| sex male | 378 (56.0%) | 722 (58.4%) | 954 (59.4%) | 456 (58.7%) | 2416 (58.4%) | 0.169 | 1027 (52.0%) | 1124 (59.9%) | 700 (56.9%) | 803 (58.7%) | 104 (58.8%) | 6134 (59.7%) | 0.453 |
| ARI category: LRI | 612 (90.7%) | 909 (73.5%) | 1191 (74.2%) | 607 (78.1%) | 3035 (73.3%) | < 0.001 | 1545 (78.2%) | 1201 (64.0%) | 773 (62.8%) | 792 (57.9%) | 133 (75.1%) | 5890 (57.3%) | 0.607 |
| codetection | 298 (44.1%) | 606 (49.0%) | 660 (41.1%) |  | 777 (18.8%) | < 0.001 |  | 118 (6.3%) | 107 (8.7%) | 129 (9.4%) |  | 177 (1.7%) |  |
| Viral pathogens |  |  |  |  |  |  |  |  |  |  |  |  |  |
|  | PCR or rapid test<sup>d</sup> |  |  |  | PCR Panel 2<sup>d</sup> |  |  |  |  |  |  |  |  |
| Patient characteristics | Flu A positive (N=6614) | Flu B positive (N=1677) | RSV positive (N=810) | p-value* | HAdV positive (N=413) | HEV positive (N=161) | MPV positive (N=121) | Codetection (N=149) | p-value* | Total all viral assays (N=42300) |  |  |  |
| age Mean (95%CI) | 4.9 (4.8, 5.0) | 5.6 (5.4, 5.8) | 1.0 (0.96, 1.1) | < 0.001 | 3.0 (2.8, 3.2) | 2.8 (2.5, 3.1) | 2.8 (2.5, 3.1) | 2.7 (2.4, 3.0) | 0.084 | 3.6 (3.5, 3.6) |  |  |  |
| sex male | 3736 (56.6%) | 979 (58.4%) | 465 (57.4%) | 0.189 | 256 (62.1%) | 92 (57.5%) | 79 (65.3%) | 88 (59.1%) | 0.620 | 24127 (57.4%) |  |  |  |
| ARI category: LRI | 604 (9.1%) | 123 (7.3%) | 685 (84.6%) | < 0.001 | 93 (22.5%) | 45 (28.0%) | 60 (49.6%) | 54 (36.2%) | < 0.001 | 8169 (19.4%) |  |  |  |
| codetection | 10 (0.2%) | 8 (0.5%) | 14 (1.7%) | < 0.001 | 98 (23.7%) | 70 (43.5%) | 53 (43.8%) |  | < 0.001 | 149 (0.4%) |  |  |  |

**(a)** Allplex Respiratory Panel 4 using real-time Polymerase Chain Reaction (PCR) of nasopharyngeal samples for the detection of 7 bacteria causing respiratory tract infections including *Bordetella parapertussis* (BPP), *Bordetella pertussis* (BP), *Chlamydomphila pneumoniae* (CP), *Haemophilus influenzae* (HI), *Legionella pneumophila* (LP), *Mycoplasma pneumoniae* (MP), *Streptococcus pneumoniae* (SP). The table shows patient characteristics for the total number of patients tested and the number of patients positive for each pathogen.
**(b)** Blood IgM serology test for *Mycoplasma pneumoniae* (MP). The table shows patient characteristics for the number of patients with IgM seropositive for *Mycoplasma pneumoniae*.
**(c)** Bacterial culture of nasopharyngeal samples. The table shows patient characteristics for the total number of patients tested and the number of patients with positive culture for each pathogen: *Haemophilus influenzae* (HI), *Streptococcus pneumoniae* (SP), *Moraxella catarrhalis* (MC).
ARI: Acute respiratory infection; LRI: Lower respiratory infection.
Codetection: $\geq 2$ pathogens were detected.
95%CI: 95% confidence interval.
\*p-values for comparison between groups are from Kruskal-Wallis test for continuous variables or Chi-square test for categorical variables.

**Figure 1.**
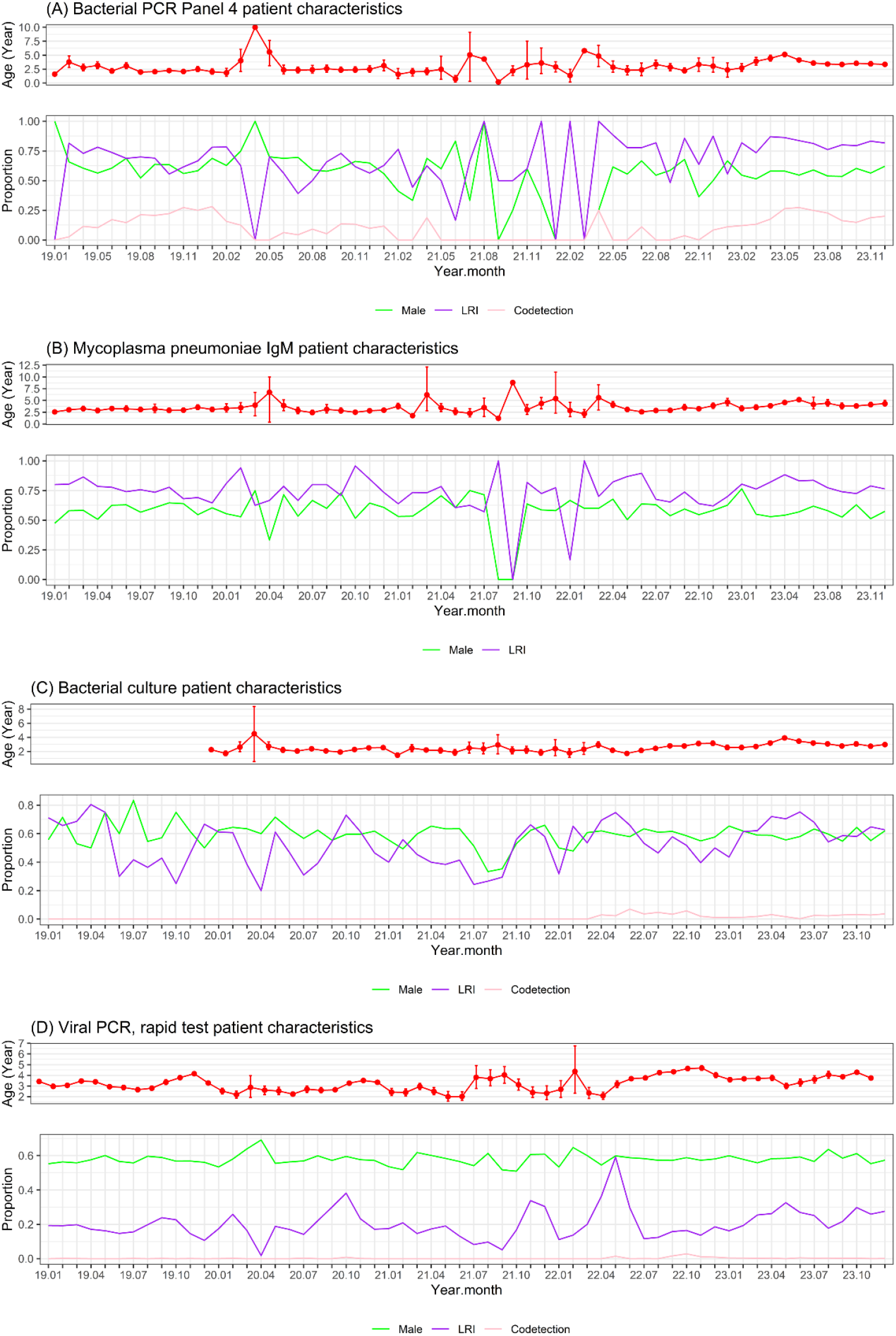
Time series plot of patient characteristics. PCR: Polymerase Chain Reaction; LRI: Lower respiratory infection. Codetection: ≥2 pathogens were detected.

Bacterial PCR Panel 4 was done for 4125 samples of 3708 patients of which LRI accounted for 73%, males accounted for 58%, the average age was 3.2 years. There were 1605 samples (39%) positive for HI, 1237 (30%) positive for SP, 675 (16%) positive for MP, and 777 (18.8%) positive for ≥2 pathogens (co-detection). Co-detection was found in 44% of those positive with MP, 49% in those positive with SP and 41% in those positive with HI. Nearly all (90%) of those positive for MP had LRI and approximately three fourths of those positive with SP or HI had LRI. The average age of the patients positive for SP or HI was less than 3 years old while the average age of those positive for MP was nearly 5 years old (p<0.001) (**Table 1, Figure 1A**). There was 1 peak of HI and SP infection in the end of 2019 before COVID-19 outbreak and a smaller peak in the end of 2020 before a close-to-zero flat period during the pandemic year 2021. A much higher peak of HI and SP infection reemerged in middle of 2023 in parallel with a newly emerged large peak of MP infection. A new small peak of CP infection also briefly emerged a few months later in middle of 2023 (**Figure 2A**).

**Figure 2.**
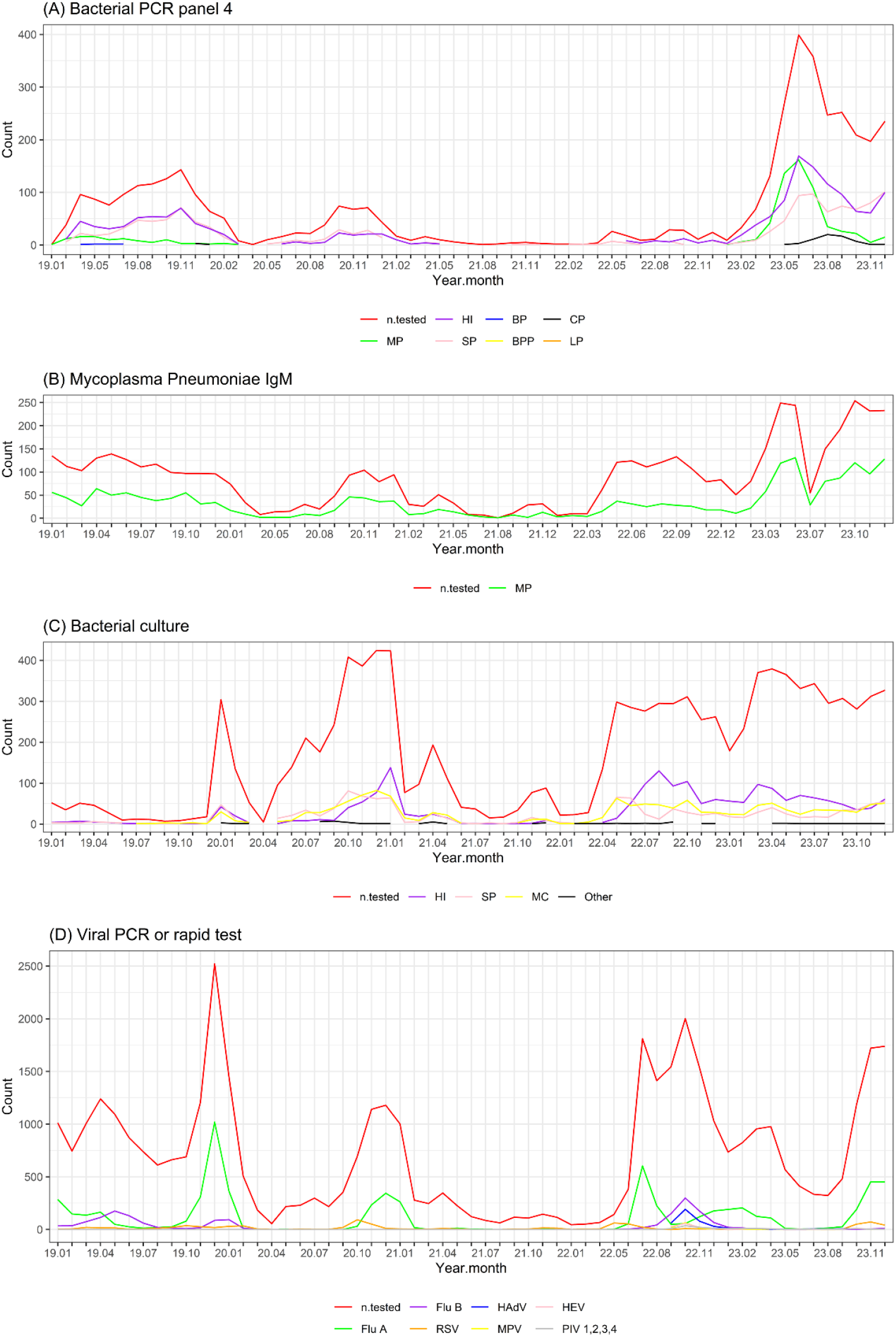
Time series plots of bacterial and viral pathogens associated with acute respiratory infections in children visited Vinmec Times City International Hospital from January 2019 to December 2023. **(A)** Allplex Respiratory Panel 4 using real-time Polymerase Chain Reaction (PCR) of nasopharyngeal samples for the detection of 7 bacteria causing respiratory tract infections including *Bordetella parapertussis* (BPP), *Bordetella pertussis* (BP), *Chlamydophila pneumoniae* (CP), *Haemophilus influenzae* (HI), *Legionella pneumophila* (LP), *Mycoplasma pneumoniae* (MP), *Streptococcus pneumoniae* (SP). The plot show the number of patients tested (n.tested) and the number of patients positive for each pathogen. **(B)** Blood IgM serology test for *Mycoplasma pneumoniae*. The plot show the number of patients tested (n.tested) and the number of patients with IgM seropositive for *Mycoplasma pneumoniae*. **(C)** Bacterial culture of nasopharyngeal samples. The plot show the number of patients tested (n.tested) and the number of patients with positive culture for each pathogen. *Haemophilus influenzae* (HI), *Streptococcus pneumoniae* (SP), *Moraxella catarrhalis* (MC) and other pathogens. **(D)** Polymerase Chain Reaction (PCR) or rapid test for *Influenza type A* (Flu A), *Influenza type B* (Flu B), *Respiratory Syncytial Virus* (RSV) and Allplex Respiratory Panel 2 using one-step real-time RT-PCR of nasopharyngeal samples for the detection of 7 viruses causing respiratory tract infections including *Human Adenovirus* (HAdV), *Enterovirus* (HEV), *Metapneumovirus* (MPV), *Parainfluenza virus* 1,2,3 4 (PIV1,2,3,4). The plot show the number of patients tested (n.tested) and the number of patients positive for each pathogen. X-axis shows year and month of the year (e.g. 19.01 means January 2019, 23.10 means October 2023). Codetection: ≥2 pathogens were detected.

There were 5049 samples of 4361 patients tested for MP IgM of which 1976 samples (39%) were serologically positive. The average age of those with seropositive MP IgM was 3.2 years old and 78% had LRI and 52% were males (**Table 1, Figure 1B**). There was no clear peak of MP IgM seropositive before and during the COVID-19 pandemic from Jan 2019 to early 2022. In middle of 2022, even though there was an increase in number of patients tested, the number of MP IgM seropositive remained low. Until March 2023, a new peak of MP IgM seropositive began to emerge in parallel with the peak of MP PCR positive in 2023 (**Figure 2B**).

Bacterial culture was done for 10280 samples of 8073 patients of which 3138 samples (31%) were positive. LRI accounted for 57%, males accounted for 59% and the average age was 2.6 years old. The most common culture positive pathogens were HI (N=1878 (18%)), SP (N=1230; 12%) and *Moraxella catarrhalis* (MC)(N=1367; 13%). Codetection of ≥2 pathogens was found in 177(1.7%). Co-detection was found in approximately 6% in those positive with HI, and 9% in those positive with SP or MC (**Table 1, Figure 1C**). There was one peak of HI, SP and MC infection in the end of 2020 to early 2021 and another large, long lasting increase starting in the middle of 2022 until the end of data period in December 2023 (**Figure 2C**).

Regarding viral pathogens, single PCR or rapid test for Flu A, Flu B, RSV or Allplex PCR Panel 2 was done for 42041 samples of 28178 patients, of which 6614 samples was positive for Flu A, 1677 positive for Flu B, 810 positive for RSV, 413 positive for HAdV, 161 positive for HEV and 121 positive for MPV. LRI accounted for <10% of those positive with Flu A or Flu B, 85% of those positive with RSV, 23% of those positive with HAdV, 28% of those positive with positive with HEV and 50% of those positive with MPV. Co-detection was common in those positive with HAdV, HEV or MPV (24% to 44%). Males accounted for approximately 60% in those positive with viral pathogens. The average age of those positive for HAdV, HEV or MPV was approximately 3 years old, the average age of those positive with RSV was 1 year old while the average age of those positive with Flu A or Flu B was approximately 5 years old (p<0.001) (**Table 1, Figure 1D**). Flu A showed a regular cycle with peaks in winter-spring months of 2019-2020, 2020-2021, 2022-2023 and end of 2023 (except a close-to-zero flat period during the COVID-19 related restriction in 2021 to early 2022). The Flu A peaks were often followed by smaller Flu B peaks a few months after. An exception was an extra early peak of Flu A in summer months of 2022 followed by a smaller peak of Flu B three months after. Another exception was a newly emerged peak of HAdV in the second half of 2022 (**Figure 2D**).

## DISCUSSION

Our analysis of microbiology assay data of nasopharyngeal samples and blood samples of children with ARI from January 2019 to December 2023 provides an overall picture of the pattern of respiratory infections other than COVID-19 and emerging pathogens before, during and after the COVID-19 pandemic in Hanoi, Vietnam.

As shown by Allplex PCR panel 4 and bacterial culture, the rebound of HI and SP was not only large in number of positive cases but also long lasting. The increase in number of HI positive cases was observed from middle of the year 2022 till the end of our data period in December 2023. About three fourths of these positive cases had LRI. This indicates that, after the COVID-19 pandemic, HI and SP have largely re-emerged and become persistent epidemic pathogens associated with LRI in children.

Although the number of patients tested for MP IgM increased since early 2022, the number of patients with seropositive MP IgM did not increase until middle of 2023. The increase in the number of patients with seropositive MP IgM was in parallel with the increase in the number of patients with positive MP PCR at the same period. This indicates that the increase was due to the real increase in the number of MP infection cases, not due to the increase in the number of patients tested. Most of the patients positive with MP had LRI and older than other patients positive with other bacterial pathogens. There was also a small newly emerged peak of CP positive cases two months after the peak of MP positive cases. These peaks were consistent with the outbreak of bacteria especially atypical bacteria associated pneumonia in 2023 late after the COVID-19 pandemic as reported in Vietnam (8), China (6) and some other countries (9).

The early rebound of Flu A and Flu B was consistent with the irregular rebound of viral ARI after the relief of COVID-19 restriction as reported in many places (4,5). The exceptional newly emerged peak of HAdV in the second half of 2022 was consistent with the unprecedented outbreak of HAdV associated ARI in children in Vietnam as previously reported (7).

The strength of our study is the relatively large number of cases with various viral and bacterial pathogens tested over 5 years that enabled us to examine the pattern and the emergence of pathogens associated with ARI in children before, during and after the COVID-19 pandemic. The main limitation of our study is that the data was from a single private hospital and might not well represent overall children population in Hanoi, Vietnam.

In brief, our data show that after the COVID-19 pandemic, HI and SP have re-emerged as epidemic pathogens associated with LRI. Flu A and Flu B have re-established regular cycles of peaks in winter-spring months after an early rebound together with an unprecedented newly HAdV emergence soon after the relief of COVID-19 restriction. Late after the COVID-19 pandemic, from middle of 2023, atypical pneumonia pathogen MP has emerged remarkably and becoming epidemic and there was also a small, brief emergence of CP infection. Our data is useful for infectious disease surveillance and public health intervention strategies.

## Data Availability

Data cannot be shared publicly because the data is the property of Vinmec Healthcare System. Data are available from Vinmec Ethics Committee (contact via corresponding author) for researchers who meet the criteria for access to data.

## Author contribution

NTH, HTTN, PMD initiated the research. HTTN, PMD, HTH, DVP obtained the data. NTH analyzed the data and drafted the initial version of the manuscript. HTTN, PMD, HTH, DVP, QNN, HTML, ANP contributed to the analysis plan and manuscript draft. All authors read and approved the manuscript.

## Ethics

This work was approved by Vinmec Ethical Committee (ref No 110/2024/CN/HDDD VMEC). Informed consent was not required for secondary analysis and report of de-identified data.

